# Insights on neonatal mortality: differences between public and private health access at São Paulo city between 2012 and 2017

**DOI:** 10.1101/2022.09.02.22278781

**Authors:** Carlos Eduardo Beluzo, Marcelo Zaccaro Zaniboni, Tiago Carvalho, Luciana Correia Alves

## Abstract

Under-five mortality is a survival measure that reflects the social, economic, and environmental conditions in which children live, including their health care, so they are reasonable measures to identify vulnerable populations. Infant Mortality, also a survival measure, refers to children dying before reaching the first year of life, and can also be disaggregated into events below 28 days, referred to as neonatal mortality, which currently concentrates the higher share of deaths. In 2020, more than 5 million under-five children died in the world, from which almost 50% occurred among newborns. The Sustainable Development Goals call for an end to preventable deaths of newborns and children under-five, but considering the current trends, many countries are not on track to achieve it. Neonatal mortality involves various biological, socioeconomic, and healthcare factors, and understanding its determinants can favor public policy funding planning and help with this problem. The increasing relevance of neonatal deaths aside from online platforms that make available health data in Brazil has enabled more precise analyses and led to a significant number of studies covering different factors, regions, and methods regarding this issue. Among the various determinants, access to private health services is an important one. In Brazil, mainly due to financial restrictions, a huge percentage of the population uses only public health services. In this context, the objective of this paper is to perform an evaluation of the NMR in the city of São Paulo - Brazil, in the period between 2012 and 2017, considering as a determinant for neonatal mortality rate if the delivery has occurred on private or on public health service sphere. Besides that, the determinant mother’s age is also included in the analysis, so it is also possible to evaluate associations between these two variables. A dataset having 8,110 neonatal deaths was analyzed and three graphics were created. The first one is used to assess information regarding the distribution of births by health service sphere and mother’s age. The second brings information on births and neonatal mortality rates combined and compared by year and by health service sphere. Finally, in the last one births and rates are also combined and compared by year, health service sphere, but now are desegregated by mother’s age group. From these results, some insights are raised and discussed, and some need further investigation to obtain more conclusive results, but the currents can already bring important knowledge about the problem.

## Introduction

The under-five mortality or child mortality is a survival measure that refers to children dying before reaching five years of age. This measure reflects the social, economic, and environmental conditions in which children (and others in society) live, including their health care, so they are good measures to identify vulnerable populations. As an important component of under-five mortality, Infant Mortality (IM) is also a survival measure that refers to children dying before reaching the first year of life, and the Infant Mortality Rate (IMR) is a Millennium Development Goals (MDG) indicator [1]. Formally, IMR is a probability of death derived from a life table and expressed as a rate per 1,000 live births. IM can be disaggregated into Neonatal Mortality (NM) and Postneonatal mortality, and Neonatal Mortality rate (NMR) is interpreted similarly to IMR, but considering events below 28 days.

Nowadays, the NM concentrates the higher share of deaths of the IM, so it has become an important object of study. According to the United Nations Inter-Agency Group for Child Mortality Estimation (UN IGME), in 2020, more than 5 million under-five children died in the world, from which almost 50% occurred among newborns, and many countries are not on track to end up under-five and newborn preventable deaths, as stated on Sustainable Development Goals (SDGs). SDGs call for an end to preventable deaths of newborns and children under-five, with all countries aiming to have an under-five mortality rate of 25 or fewer, and an NMR of 12 or fewer by 1,000 births alive, by 2030, but considering the current trends, 54 countries will not meet the target by 2030, and 61 will miss the neonatal mortality target. [2].

The graphic in **Figure 1** was extracted from the “*Levels and trends in child mortality*” website, a resource provided by United Nations Inter-Agency Group for Child Mortality Estimation (UN IGME). It depicts the NMR by region in the world in 1990, 2000, and 2020, and as can be seen, in all regions the rate is declining, but some regions may not achieve the target for 2030, represented in the graphic by the green line. Latin America and the Caribbean are on track to achieve the target, although it’s known that there is still much work to be done in some countries specifically.

**Figure 1.**
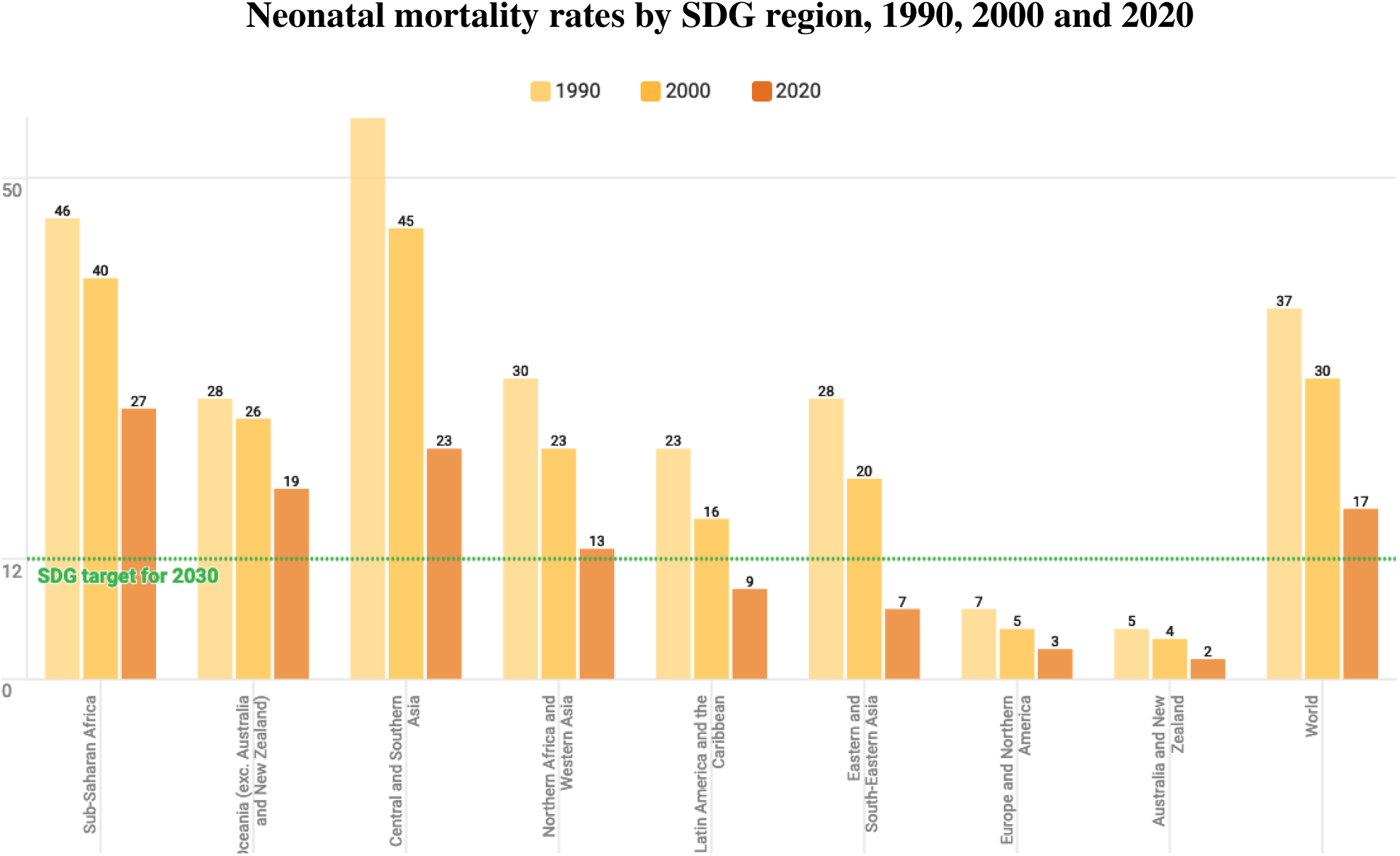
NMR by SDG region, 1990, 2000, and 2020. (**Source**: Levels and trends in child mortality - UN IGME, 2021).

In Brazil, IMR has undergone a continual decrease in recent decades, mainly due to improvements in sanitary conditions and the reduction in Postneonatal which has declined from 51% in 1990 to 38% in 2015 [4]. NMR has also been following a declining trend, mainly due to favorable changes in factors related to pregnancy and childbirth, despite the increase in its shared proportion [3]. The IMR and NMR in Brazil in the last decades are depicted in **Figures 2** and **3** respectively.

**Figure 2.**
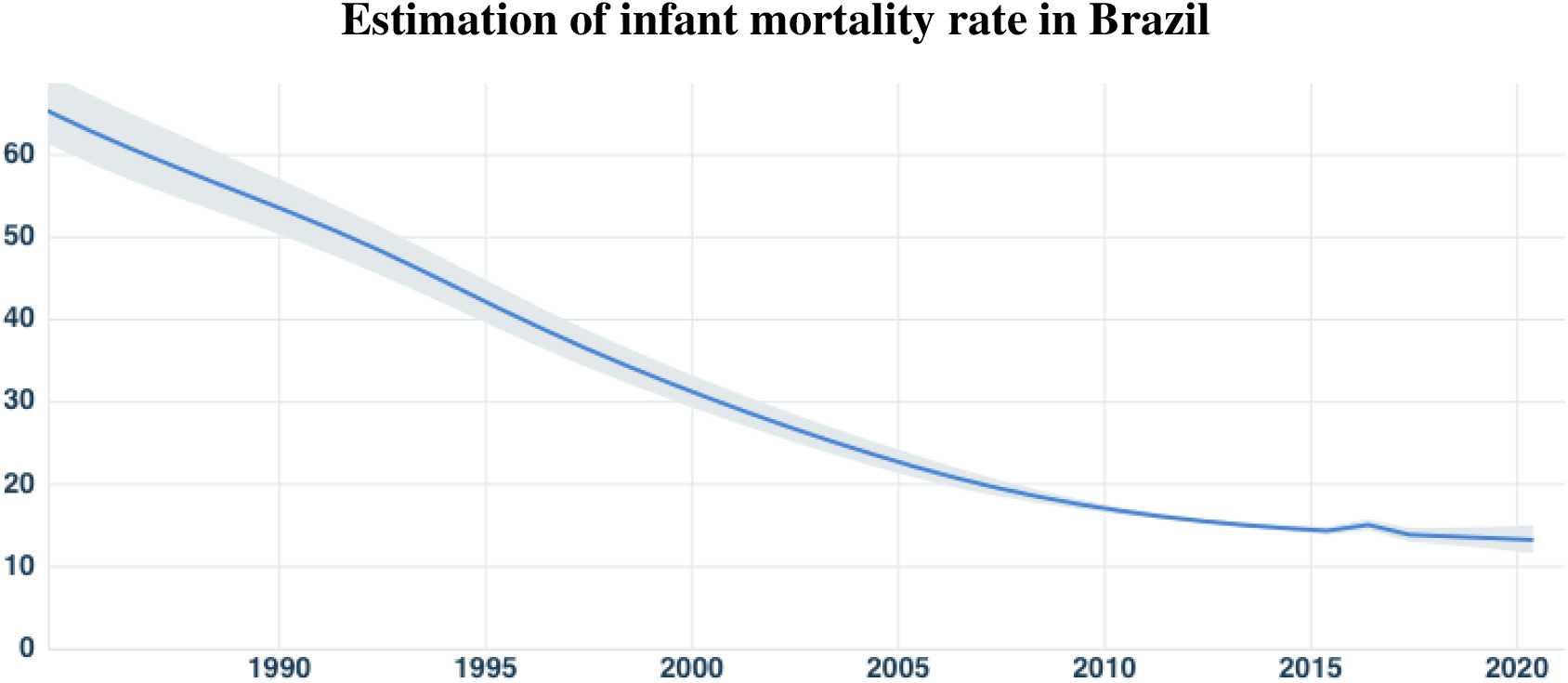
Estimation of IMR in Brazil in the last decades. (**Source**: United Nations Inter-agency Group for Child Mortality Estimation).

**Figure 3.**
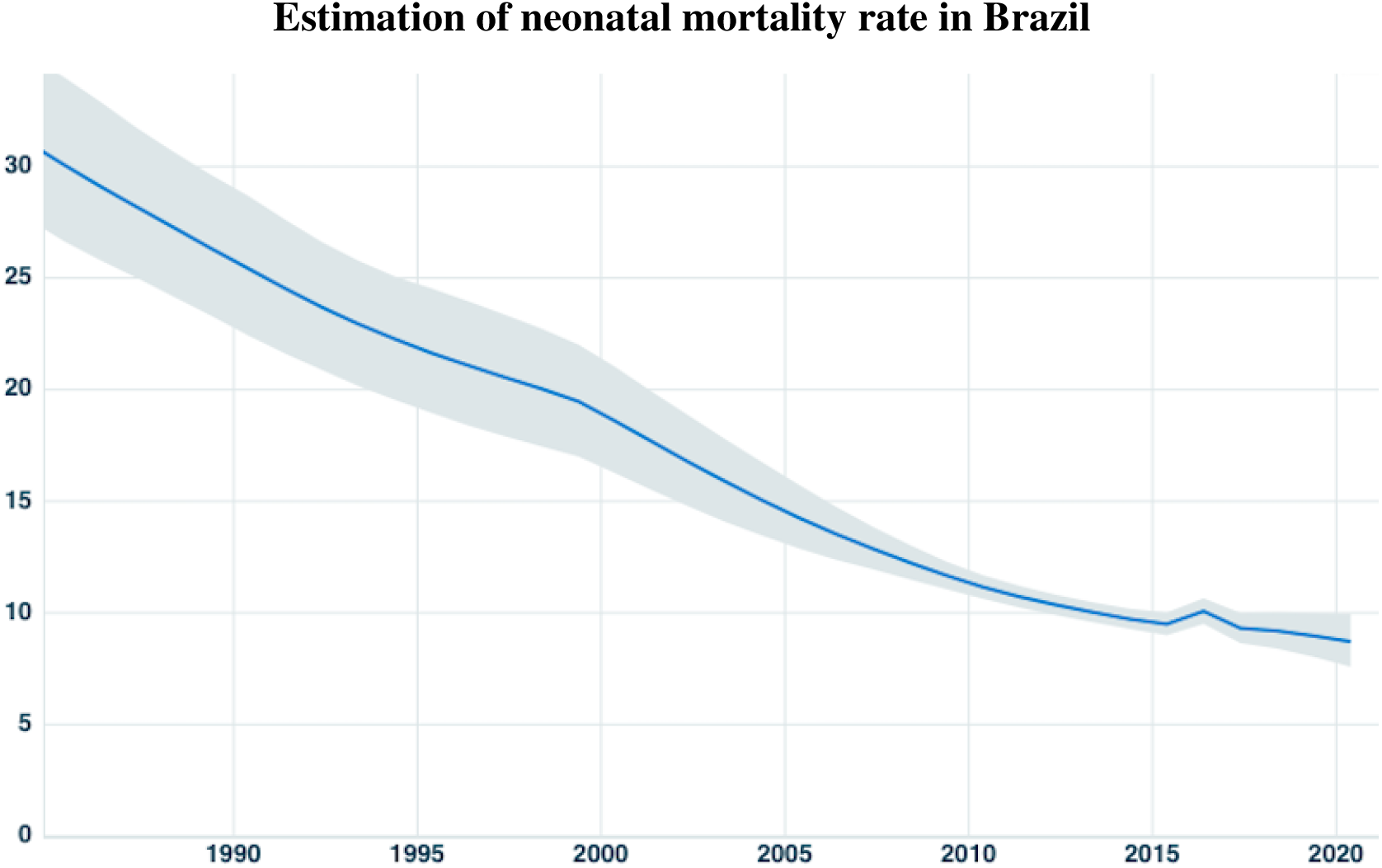
Estimation of NMR in Brazil in the last decades. (**Source**: United Nations Inter-agency Group for Child Mortality Estimation).

Understanding NM determinants can favor public policy funding planning. It involves various biological, socioeconomic, and healthcare factors and has become the biggest challenge in fighting IM. The increasing relevance of neonatal deaths on IM aside from online platforms that make available health data in Brazil, such as the Department of Brazilian Health Ministry (DATASUS) responsible for collecting, processing, and disseminating public health data, have enabled more precise analyses and led to a significant number of studies covering different factors, regions, and methods regarding this issue [2].

Among the various determinants of NM, the access to private health services, which in general offers faster medical care and consequently has faster diagnosis and intervention than in the public health sphere, is very restrictive in Brazil, mainly due to financial restrictions, so a huge percentage of the Brazilian population uses the public health service. According to data from the National Health Survey (Pesquisa Nacional de Saúde - PNS), in 2019 in Brazil, less than 30% of the population has an insurance plan, and almost 50% of the population in the city of São Paulo has some health insurance [5]. In this context, the objective of this paper is to perform an evaluation of the NMR in the city of São Paulo - Brazil, in the period between 2012 and 2017, considering as a determinant for NMR if the delivery has occurred on private or on public health service sphere. Besides that, the determinant mother’s age is also included in the analysis, so it is also possible to evaluate associations between these two variables.

## Material and Methods

The data came from Mortality Information System (SIM - Sistema de Informação de Mortalidade) and the National Information System on Live Births (SINASC – Sistema de Informação de Nascidos Vivos), both from DATASUS (Health Informatics Department of the Brazilian Ministry of Health). From there, it was extracted the dataset used in this paper, which has been built by selecting only records from São Paulo city for the period of 2012 to 2017. Data from São Paulo city has high-level quality. Although São Paulo has one of the lowest levels of neonatal mortality rates in Brazil, these events occurred in heterogeneous ways with smaller or even lower reductions in the most vulnerable populations, as the reflection of unfavorable life conditions of the population, healthcare, and socioeconomic inequalities [6].

SINASC is fed using the Live Birth Statement (DNV - Declaração de Nascido Vivo). It comprises information about demographic and epidemiological data for the infant, mother, prenatal care, and childbirth. On the other hand, SIM has the main goal of supporting the collection, storage, and management process of death records in Brazil, and was used to label records where the death happened until 28 days of life on SINASC, using DNV, which is a common field in both systems, as an association key [7].

The methodology used to create this dataset was the same used to build the **SPNeodeath** dataset [8], but in the current paper, only the features mother’s age and birth sphere (private or public health) were selected. According to data from SIM, there were 13,309 infant deaths (died before the first year of life) in the period between 2012 to 2017, in the municipality of São Paulo. Of these 8,180 refers to neonatal deaths. After data preprocessing tasks, including dealing with missing data, the dataset used in this paper has left with 8,110 records of neonatal deaths that occurred in the city of São Paulo between 2012 to 2017.

For analysis intent, three graphics were created using the neonatal deaths data, and are presented in the results section. The first one is used to evaluate information regarding the distribution of births by health service sphere and mother’s age. The second brings information on births and NMR combined and compared by year and by health service sphere. Finally, in the last one births and NMR are also combined and compared by year, health service sphere, but now are desegregated by mother’s age group.

## Results and Discussion

One relevant piece of information we can infer from the data is that *“as older as the mother is, the births happen more in the private sphere”*. This can be seen in **Figure 4**, which brings the distribution of births by health service sphere and by mother’s age between 2012 and 2017 in São Paulo city. Part of this trend is generally associated with the fact that older women have already been inserted into the labor market, and then have financial resources to pay for private health services. Otherwise, young mothers use more public health services. Another piece of information depicted in this graphic is that at about the age of 21, the number of births in the public sphere starts to decay, and about the age of 28 is almost balanced between the two spheres.

**Figure 4.**
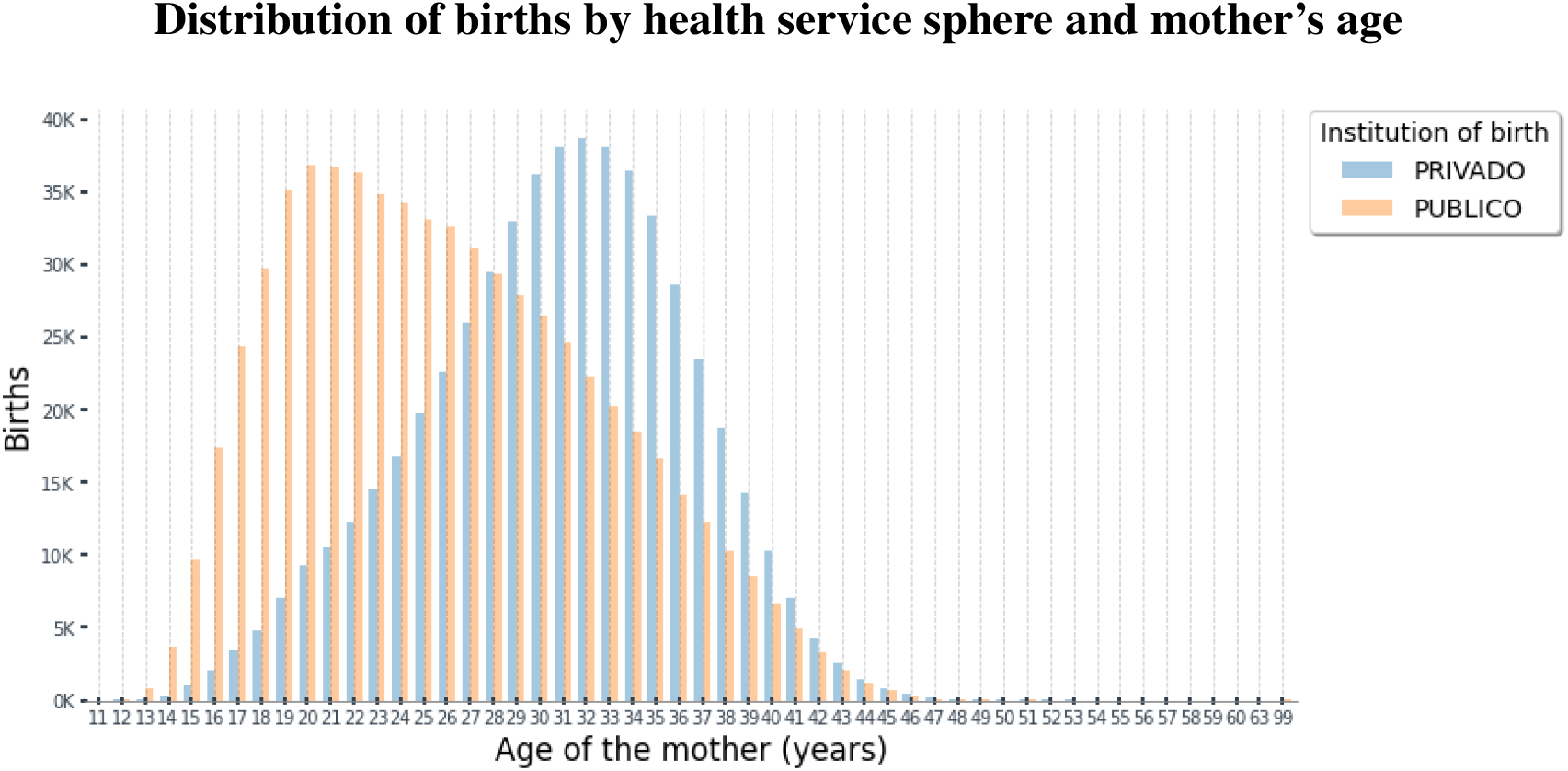
Distribution of births by health service sphere and mother’s age between 2012 and 2017 in São Paulo city. (**Source**: SIM and SINASC).

The data in **Figure 5**, brings important insights into NMR between in public and spheres. The bars in the graphic show the total absolute number of births (in the region and the period subject of this paper) for public and private health services separated, and two trend lines representing the NMR on each one of the health service spheres. From the graphic, it is possible to observe that the NMR is consistently higher in public than in private institutions. This is very important information that requires attention and deeper analysis and also further information is needed for better understanding and causes identification, but *the NMR is almost twice times higher on the public than on private services* along all the periods evaluated.

**Figure 5.**
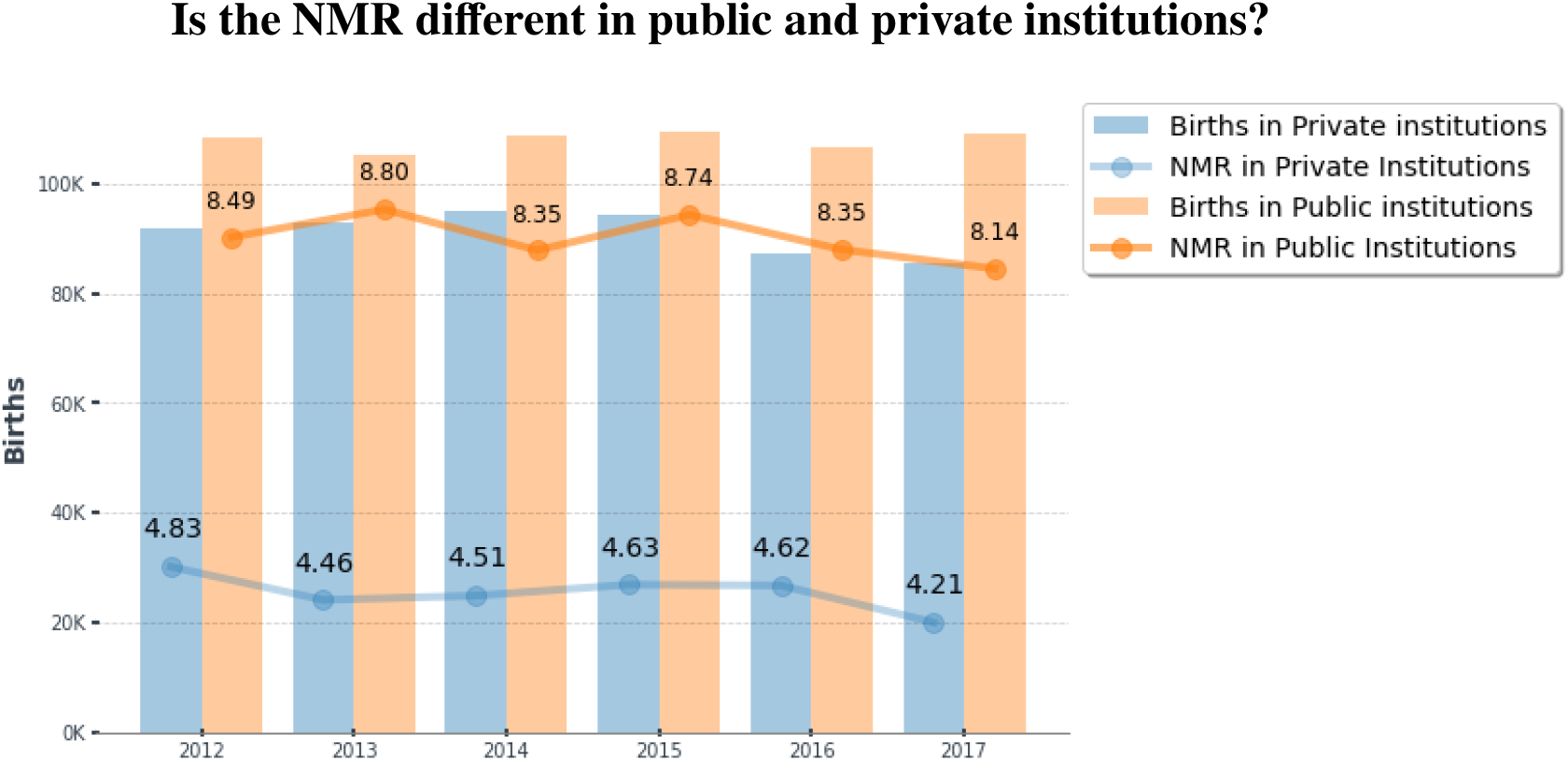
Births and NMR by health service sphere between 2012 and 2017 in São Paulo city. (**Source**: SIM and SINASC).

In this way to better understand this fact, **Figure 6** brings similar information segregated by mother’s age group, in an attempt to answer the question: *By mother’s age group, is the NMR different in public and private institutions?* From this graphic, it is important to note that in young mothers the difference in the NMR between the two spheres is lower than in mothers of older ages, and a possible reason is an effect of the knowing fact that *children of younger mothers have higher survival probabilities*.

**Figure 6.**
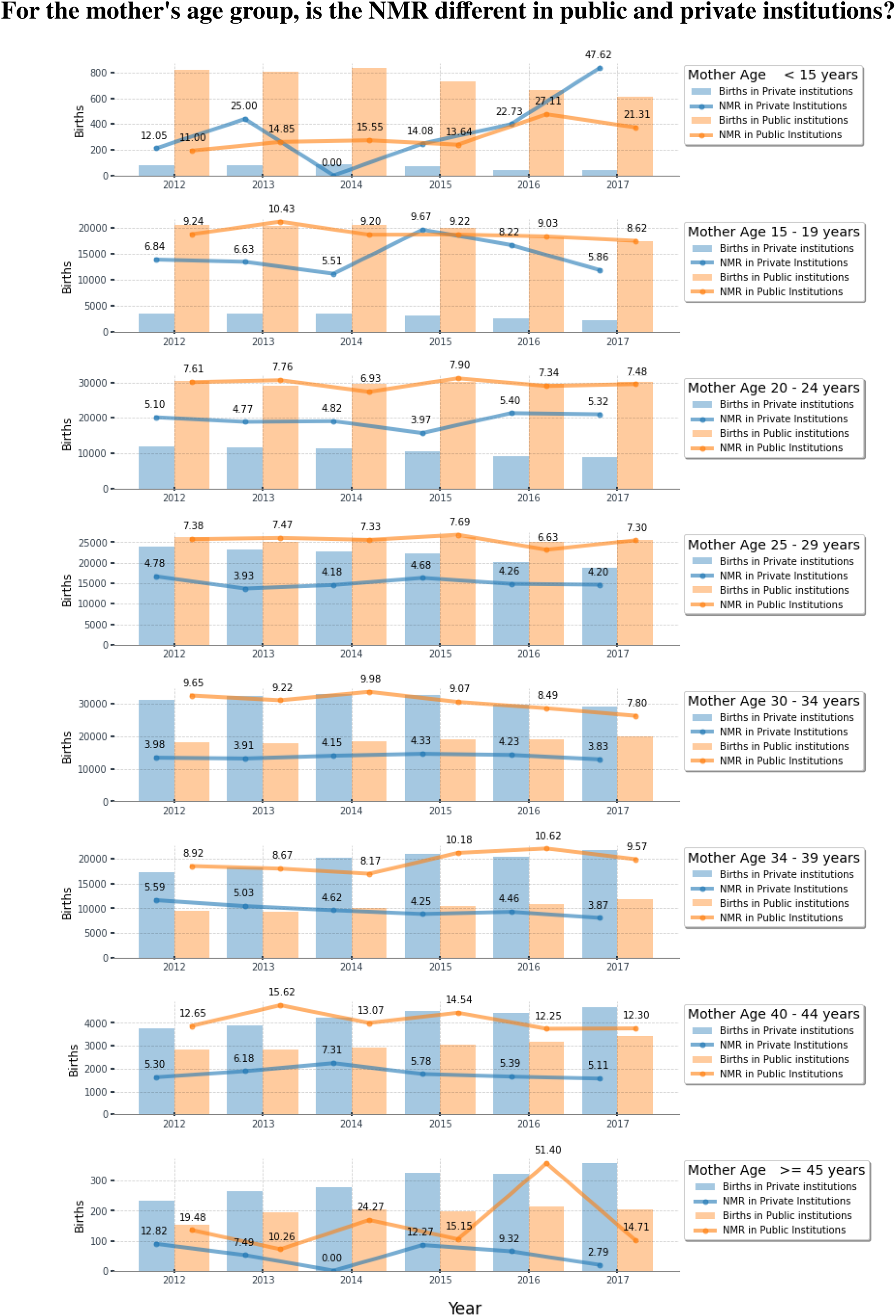
Births and NMR by health service sphere and mother’s age group between 2012 and 2017 in São Paulo city. (**Source**: SIM and SINASC).

Many insights can also be inferred from this data, and a comparison with results from other studies is being held. The current study intends to advance from this preliminary result. It is necessary to explore these relationships through Poisson regression models in order to obtain more conclusive results.

## Data Availability

All data produced in the present study are available upon reasonable request to the authors.

https://doi.org/10.1016/j.dib.2020.106093

https://opendatasus.saude.gov.br/dataset/sim-2020-2021

https://opendatasus.saude.gov.br/dataset/sistema-de-informacao-sobre-nascidos-vivos-sinasc-1996-a-2020

## Notes

### Competing Interest Statement

The authors have declared no competing interest.

### Funding Statement

This research was supported by Bill & Melinda Gates Foundation (#OPP1201970) and Ministry of Health of Brazil, through the National Council for Scientific and Technological Development (CNPq) (#443774/2018-8). It was also supported by NVIDIA, that donated a GPU XP Titan used by the research team.

### Author Declarations

Ethics committee/IRB of Brazilian Responsible Ethics Committee, Institution #5473 - Instituto Federal de Educação, Ciencia e Tecnologia de São Paulo - IFSP, SUZANO/SP - Brazil, gave ethical approval for this work under approval number #3.193.057. The approval can be validated at https://plataformabrasil.saude.gov.br using project title: "Plataforma de Apoio à Decisão para Políticas Públicas de Saúde Gestacional Baseada em Técnicas de Visualização de Informaçõ es e Aprendizado de Máquina".

